# Sociodemographic inequalities in onset, mortality and prognosis among patients developing diabetic foot ulcers: a flexible parametric analysis

**DOI:** 10.64898/2026.07.06.26355671

**Authors:** Ian Farr, Jincy James, Timothy Howcroft, Moi Hoon Yap, Neil D. Reeves, Pappachan M Joseph, Vishnu Chandrabalan

## Abstract

**Aim:** Inequalities in diabetic foot ulcer (DFU) outcomes are driven by several factors including sociodemographic factors. This study examined the intersectional risks of ethnicity, sex, and deprivation on DFU progression, which prior research often evaluated in isolation.

**Methods:** A retrospective cohort study (2009–2024) of 2,125 patients at Lancashire Teaching Hospitals Trust utilized flexible parametric survival modelling. Models assessed DFU onset, overall mortality, and post-clinic prognostic survival, adjusting for demographics and comorbidities.

**Results:** The most deprived patients presented significantly younger (median 64 vs. 73 years). Male sex accelerated DFU onset (HR: 1.24) and increased overall mortality risk (HR: 1.14). Black patients presented older with higher comorbidity burdens but paradoxically exhibited lower overall mortality risk (HR: 0.49). Deprivation heavily impacted life expectancy as the most deprived group showed higher mortality rates (HR: 0.73) and reduced 5-year prognostic survival (48.7% vs. 59.1%). Presence of comorbidities linearly increased overall mortality risk. Furthermore, severe deprivation caused greater overall life-years lost in men (4.0) than women (2.5).

**Conclusions:** Patient outcomes with DFU are heavily influenced by cumulative demographic and socioeconomic factors. Effective management requires accessible, holistic care that actively accommodates these complex biosocial-economic realities.

## Introduction

In the United Kingdom, the direct costs of managing diabetic foot ulcers (DFU) consume nearly 1% of the National Health Service (NHS) budget, exceeding the costs of care for breast, prostate, and lung cancers combined (Kerr et al., 2019). The lifetime risk for a person with diabetes developing a DFU is estimated between 19% and 34% (Edmonds et al., 2021). The DFU acts as a sentinel event in terms of increased mortality risk, because of the presence of complications including atherosclerosis and neuropathy (Boulton et al., 2020, Jean-Baptiste et al., 2025). Underscoring this, the five-year mortality rate following DFU onset is reported to be between 50% and 70%, and those with diabetes who develop a foot ulcer are twice as likely to die within ten years compared to those without ulcers (Rubio et al., 2020). However, this burden is not distributed equally, and a clinically stratified risk profile is evidenced across ethnicity, deprivation, and sex (McDermott et al., 2023).

In terms of ethnicity, although South Asian populations have higher body fat to BMI ratio and higher type 2 diabetes prevalence at younger ages, they exhibit a lower risk of diabetic foot ulcers (DFUs), with risk levels up to one-third that of White Europeans (Abbott et al., 2005). This is attributed to lower smoking rates, lower incidence of foot deformities and reduced risk for peripheral arterial disease. Conversely, Black Caribbean and African cohorts experience significantly higher rates of infection and further complication (Chaturvedi et al., 2001; Holman et al., 2012). More than just an isolated factor, these disparities highlight the potential for ethnicity, socioeconomic deprivation, and systemic healthcare access to intersect and drive global health inequalities.

Socioeconomic deprivation functions as an amplifier of clinical risk as it influences DFU outcomes through multiple mechanisms including: financial barriers to appropriate footwear, occupational demands, nutritional deficits, and residence in areas with poor walkability (Bonnet & Sultan, 2022; Fallon et al., 2022). Consequently, DFU-related amputation rates in low-income neighbourhoods can be double those in high-income areas (Bilal et al., 2023; Fallon et al., 2022). Recent analyses of deprivation indicate that the risk of death in DFU patients increases by approximately 14% for every quintile increase in deprivation (Bilal et al., 2023).

Historically, male sex has been the predominant risk factor for ulceration due to apparent occupational and cardiovascular profiles (Vanherwegen et al., 2023). However, the rising prevalence of young-onset type 2 diabetes is shifting this established demographic landscape (Dhadse et al., 2024). A notable sex-survival discordance has also emerged. Women exhibit a lower incidence of ulcers yet frequently face inferior post-amputation survival outcomes (Seghieri et al., 2023). The drivers of this survival disparity remain unclear, although it is noted that female patients often present at a significantly more advanced age than their male counterparts. The confounding factors of age and socioeconomic status currently obscure the independent relationship between sex and mortality.

These social drivers of health inequalities may compound biological risk. The weathering hypothesis suggests that chronic exposure to socioeconomic marginalization induces cumulative physiological wear and tear, effectively accelerating biological aging and precipitating clinical complications earlier in the life course (Geronimus et al., 2006). In the context of diabetic foot disease, this weathering effect likely manifests as a premature shift in the age of onset among deprived cohorts, where the allostatic load of poverty mimics the clinical profile of advanced chronological age.

Synthesizing these distinct strands of inequality into a comprehensive understanding of DFU risk remains a challenge. Current epidemiological data often examines these risk factors in isolation or within non-comparable datasets, limiting the ability to adjust for confounding variables rigorously or for comparison across studies. Moreover, while the relative risk of these biosocial determinants is comparatively well-established, the absolute difference in life expectancy remains unclear. Demonstrating the absolute survival outcomes can provide clinically useful and easily interpretable information. Therefore, a robust data framework is required to translate these complex risk profiles.

The Observational Medical Outcomes Partnership (OMOP) Common Data Model (CDM) standardizes Electronic Health Records (EHR) into a common format, ensuring consistent definitions of exposures and outcomes across healthcare systems. By leveraging this standardized infrastructure for reproducible epidemiology, this study seeks to quantify the absolute impact of the potential intersectional biosocial health inequities of ethnicity, deprivation, and sex on the age of DFU onset, prognostic mortality, and age at death.

## Methods

### Study Design and Data Source

This was a retrospective cohort study using Lancashire Teaching Hospitals NHS Foundation Trust (LTHTR) OMOP–CDM data version 5.4. The study period was defined from 2009-06-01 to 2024-03-14. Source systems contributing to the health data processing were the diabetic foot clinic clinical photo repository. OMOP event and patient history data depended on the Patient Administration System (PAS) source data. The publicly available Index of Multiple Deprivation 2019 (IMD) was used to inform the sociodemographic analysis (UK GOV, 2020). Ethical approval for this study was obtained from the NHS Research Ethics Committee (REC reference: 15/NW/0539; IRAS project ID: 159863).

### Study Population

The cohort comprised 2,125 adult patients older than 18 years age who had attended the LTHTR foot clinic with an active DFU and had a DFU photograph entry for that event.

Patients were excluded if they had missing demographic data essential for the primary analysis (year of birth) or inconsistent data quality flags (e.g., death date recorded prior to the index event). A total of 25 patients were removed during the data cleaning phase due to such inconsistencies.

### Outcomes and predictors

Socioeconomic deprivation: assessed using the IMD derived from the Lower Super Output Area (LSOA) of residence (UK GOV, 2020). To mitigate endogeneity bias, the health domain was excluded from the IMD calculation, and ranks were recalculated specifically for this cohort (Mohammed et al., 2024).

Deprivation: categorized into quintiles, ranging from Quintile 1 (most deprived) to Quintile 5 (least deprived). Ethnicity was self-reported and mapped to standard OMOP concept IDs, and sex was extracted from the clinical register. Comorbidity burden was summarized using the Charlson Comorbidity Index (CCI), a weighted score of 17 chronic conditions (Gasparini, 2018).

Death: identified through a combination of LTHTR administrative records and GP patient death notification system.

Outcomes: 1) Age at onset was modelled as the time from birth to the index date (first specialist clinic visit). 2) Age at death (overall mortality) was modelled from birth to death using a left-truncated approach to account for delayed entry at the age of DFU presentation. 3) Clinical prognosis was defined as the time from the index DFU clinic visit to death.

### Statistical Analysis

Continuous variables (e.g., age at presentation) were summarized using means (SD) or medians (IQR) depending on normality. Categorical variables were reported as counts and percentages. Group differences in baseline characteristics were assessed using Kruskal-Wallis tests for continuous variables and Chi-squared tests for categorical variables.

To overcome the limitations of the Cox Proportional Hazards (CPH) model, specifically the limitation to estimate absolute survival and time-varying effects, we used Royston-Parmar Flexible Parametric Survival Models (FPM). The baseline hazard function was modelled using restricted cubic splines on the log-cumulative hazard scale. Complexity of the hazard function was first tested at 3 degrees of freedom for all models to ensure clinical interpretability and model stability unless Cox-Snell residuals showed poor goodness-of-fit.

The proportional hazards assumption was formally tested for ethnicity, sex, and deprivation. Where the assumption was violated, we incorporated Time-Varying Covariates (TVC) using spline-based interactions with time. Optimal TVC complexity (degrees of freedom) was determined by minimizing the Akaike Information Criterion (AIC).

To quantify the absolute impact of health inequities, we calculated the Restricted Mean Survival Time (RMST). This allowed us to translate relative Hazard Ratios into years of life gained or lost across specific time periods (e.g. age 40 to 90). Absolute survival probabilities were predicted to visualize the difference in the age of DFU onset for deprived groups. Adjusted Hazard Ratios (aHR) and 95% Confidence Intervals (CI) were calculated using the Delta method to ensure numerical stability.

All statistical analyses were performed using R (Version 4.3.0).

## Results

The final analytic cohort comprised 2,125 patients. The population was predominantly male (n= 1,399, 66%; female n = 726, 34%) with a median age at first DFU presentation of 69 years (SD = 14.8; female age at first clinic = 73, SD=16.3). These baseline sex differences are presented in table 1. Baseline characteristics revealed significant socioeconomic gradient as patients in the most deprived quintile (Q1) presented at a significantly younger age (median 69 vs. 71 years for Q5, p=0.008) and carried a higher burden of severe comorbidities compared to the least deprived group. Due to low numbers in some groups, ethnic categories were further pooled as “White” (n=1986), “Asian” (n=76), “Black” (n=23), and “Unknown” (n=40). Baseline comparisons by deprivation and ethnicity are presented in supplementary tables S1 and S2.

**Table 1:** Baseline characteristics by sex.

| Characteristic | Male | Female | p-value<sup>1</sup> |
| --- | --- | --- | --- |
|  | N = 1,399 | N = 726 |  |
| Age at Presentation<br>(Years)<sup>2</sup> | 69 (59,<br>79) | 73 (60,<br>82) | <0.001 |
| Deprivation Quintile, n<br>(%) |  |  | 0.24 |
| Q1 - Most Deprived<br>(In cohort) | 236 (17) | 108 (15) |  |
| Q2 | 239 (17) | 112 (15) |  |
| Q3 | 217 (16) | 132 (18) |  |
| Q4 | 226 (16) | 128 (18) |  |
| Q5 - Least Deprived<br>(In cohort) | 223 (16) | 128 (18) |  |
| Unknown | 258 (18) | 118 (16) |  |
| Ethnicity, n (%) |  |  | 0.74 |
| Asian | 52 (3.7) | 24 (3.3) |  |
| Black | 17 (1.2) | 6 (0.8) |  |
| White | 1,302<br>(93) | 684 (94) |  |
| No matching concept | 28 (2.0) | 12 (1.7) |  |
| Body Mass Index (kg/m<sup>2</sup>)<sup>2</sup> | 27 (22,<br>32) | 26 (20,<br>32) | 0.072 |
| Unknown | 680 | 392 |  |
| Charlson Comorbidity<br>Index<sup>2</sup> | 3.00<br>(1.00,<br>4.00) | 3.00<br>(1.00,<br>5.00) | 0.7 |
| <sup>1</sup> Wilcoxon rank sum test; Pearson's Chi-squared test; <sup>2</sup> Median<br>(Q1, Q3) |  |  |  |

Baseline characteristics differed significantly by deprivation level. Patients in the most deprived quintile (Q1) were significantly younger at presentation (median 64 vs. 73 years in Q5; p<0.001) and more likely to be Asian ethnicity (13% in Q1 vs. 2.0% in Q5; p<0.001). Notably, the Black cohort presented with a high baseline comorbidity burden (median CCI 3.00, Q3 5.00).

### Onset

Modelling the cumulative probability of developing a DFU over the life course revealed a time-varying effect on the age of onset (p=0.03) associated with deprivation. This onset disparity was most pronounced in middle age, with survival curves for the most deprived quintile diverging sharply from the least deprived by age 45–50 (Figure 1a).

**Figure 1a:**
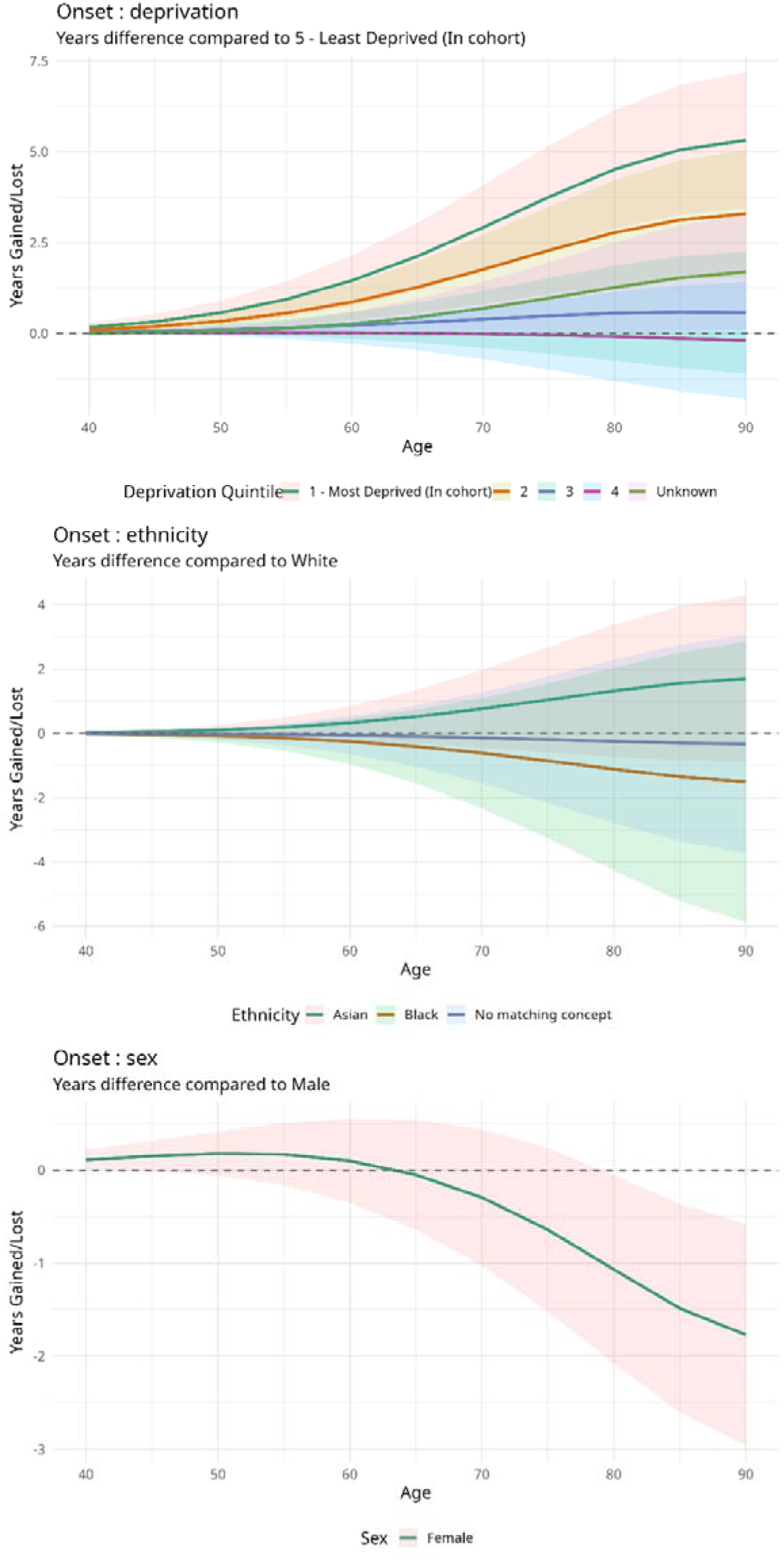
Onset RMST. Note: Positive values on the Y-axis indicate greater years lost relative to the reference group

These absolute time differences are illustrated in Figure 1a using Restricted Mean Survival Time (RMST) estimates of differences in DFU-free survival across deprivation, ethnicity, and sex. Panel A shows that among men, the socioeconomic disparity widens progressively with age, with the most deprived quintile increasingly diverging from the least deprived reference group (Quintile5), particularly after later adulthood. Panel B demonstrates smaller but distinct ethnic differences in DFU-free survival relative to White patients, with Asian patients showing increasing divergence over time. In Panel C, females display an age-dependent trajectory relative to males, with differences becoming more pronounced in older age groups rather than remaining constant. The deprivation gradient is substantially larger than the ethnicity-related differences, suggesting that socioeconomic deprivation is strongly associated with earlier DFU onset over the life course. These results suggest that deprivation acts as a powerful, time-dependent accelerator of DFU onset, particularly among men.

These gaps represent a substantial difference in absolute disease-free survival. Our estimations showed that at age 70, the probability of remaining DFU-free was 13.4% higher for the least deprived quintile (57.0%) than for the most deprived (43.6%). The wider confidence ribbons at the tails of the age distribution reflect data sparsity among the youngest and oldest patients rather than poor model fit.

The interaction between ethnicity and deprivation was not significant (p=0.39), indicating that the disparities associated with high deprivation are experienced similarly across all ethnic groups. In the multivariable model (Table 2), male sex was associated with a significant acceleration of disease onset (HR 1.24, 95% CI 1.13–1.36, p<0.001).

**Table 2:** Flexible parametric survival model estimates for onset, overall mortality, and prognosis.

|  | **Age at first clinic**<br>**visit (onset)** |  | **Age at death**<br>**(overall mortality)** |  | **Prognosis** |  |
| --- | --- | --- | --- | --- | --- | --- |
| **Predictor** | **HR (LCI-**<br>**UCI)** | **p-**<br>**value** | **HR (LCI-**<br>**UCI)** | **p-**<br>**value** | **HR (LCI-**<br>**UCI)** | **p-**<br>**value** |
| **Male sex (ref:**<br>**Female)** | 1.24 (1.17,<br>1.3) | <0.001 | 1.14 (1.06,<br>1.22) | 0.043 | 1.11 (1.03,<br>1.19) | 0.095 |
| **Deprivation Q2 (ref:**<br>**Q1)** | 0.85 (0.77,<br>0.95) | 0.039 | 0.85 (0.73,<br>0.97) | 0.106 | 0.88 (0.76,<br>1) | 0.209 |
| **Deprivation Q3** | 0.68 (0.61,<br>0.76) | <0.001 | 0.83 (0.72,<br>0.95) | 0.073 | 0.82 (0.71,<br>0.94) | 0.065 |
| **Deprivation Q4** | 0.65 (0.59,<br>0.72) | <0.001 | 0.76 (0.66,<br>0.87) | 0.009 | 0.76 (0.66,<br>0.87) | 0.009 |
| **Deprivation Q5**<br>**(Least deprived)** | 0.68 (0.61,<br>0.75) | <0.001 | 0.73 (0.63,<br>0.84) | 0.003 | 0.73 (0.63,<br>0.84) | 0.003 |
| **Deprivation**<br>**(Unknown)** | 0.8 (0.73,<br>0.89) | 0.004 | 0.84 (0.72,<br>0.96) | 0.09 | 0.83 (0.72,<br>0.95) | 0.082 |
| **Ethnicity: Asian**<br>**(ref: White)** | 1.17 (0.93,<br>1.45) | 0.194 | 1.05 (0.75,<br>1.42) | 0.787 | 1.03 (0.74,<br>1.4) | 0.846 |
| **Ethnicity: Black**<br>**(ref: White)** | 0.88 (0.57,<br>1.29) | 0.541 | 0.49 (0.25,<br>0.86) | 0.025 | 0.51 (0.25,<br>0.89) | 0.033 |
| **Ethnicity: No matching concept (ref: White)** | 0.97 (0.7, 1.3) | 0.856 | 1.09 (0.61, 1.77) | 0.747 | 1.08 (0.61, 1.75) | 0.779 |
| **CCI** | 0.98 (0.97, 0.99) | 0.009 | 1.08 (1.06, 1.09) | <0.001 | 1.07 (1.06, 1.08) | <0.001 |
| **Age at presentation** | - | - | - | - | 1.05 (1.05, 1.05) | <0.001 |

*Note: for overall mortality, age at presentation was included in the model as a continuous variable using restricted cubic splines (3 degrees of freedom) to account for non-linear effects. - : age at presentation was not modelled in for these outcomes.*

### Age at death

In the analysis of overall survival, an ethnicity paradox was observed regarding mortality. While unadjusted trends suggested lower survival for ethnic minority groups, the adjusted model indicated a protective effect for the Black cohort (HR 0.47, 95% CI 0.25–0.88, p=0.018). This divergence suggests negative confounding driven by disease severity as the Black cohort presented with a significantly higher comorbidity burden than the White cohort (Table 2). Once the model adjusted for this disparity, a latent survival advantage was revealed.

To address numerical instability and convergence failure when estimating a single hazard ratio for sex, we utilized a sex-stratified approach. Independent models were fitted for males and females. For males, a likelihood ratio test confirmed that a time-varying model was superior to a proportional hazards model X^2^ = 16.4, df = 4, p = 0.0025). As this test was not significant for females, a standard proportional hazards model was maintained for that group.

Deprivation remained a persistent driver of mortality. Patients in the most deprived quintile experienced a tangible loss of life expectancy compared with the least deprived (HR 0.73, 95% CI 0.63–0.84, p=0.003). Comorbidity burden acted as a linear predictor; each single-point increase in the CCI was associated with an 8% increase in the hazard of death (HR 1.08, 95% CI 1.06–1.09, p <0.001). Notably, for men, this effect was age-dependent (time-varying), whereas for women it was a constant proportional hazard. Presenting the hazard ratios for deprivation as age-dependent functions shows high deprivation resulted in 4 years of life lost for men, compared to 2.5 years for women.

Figure 1b unpacks this picture, stratified by sex. Panel A shows differences in RMST for the male cohort by deprivation quintile, using Quintile 5 (least deprived) as the reference group (dashed zero line). The most deprived male cohort demonstrates the largest divergence from the reference group across age, with the gap widening progressively over time. This indicates substantial inequality in expected survival between deprivation groups among men.

**Figure 1b:**
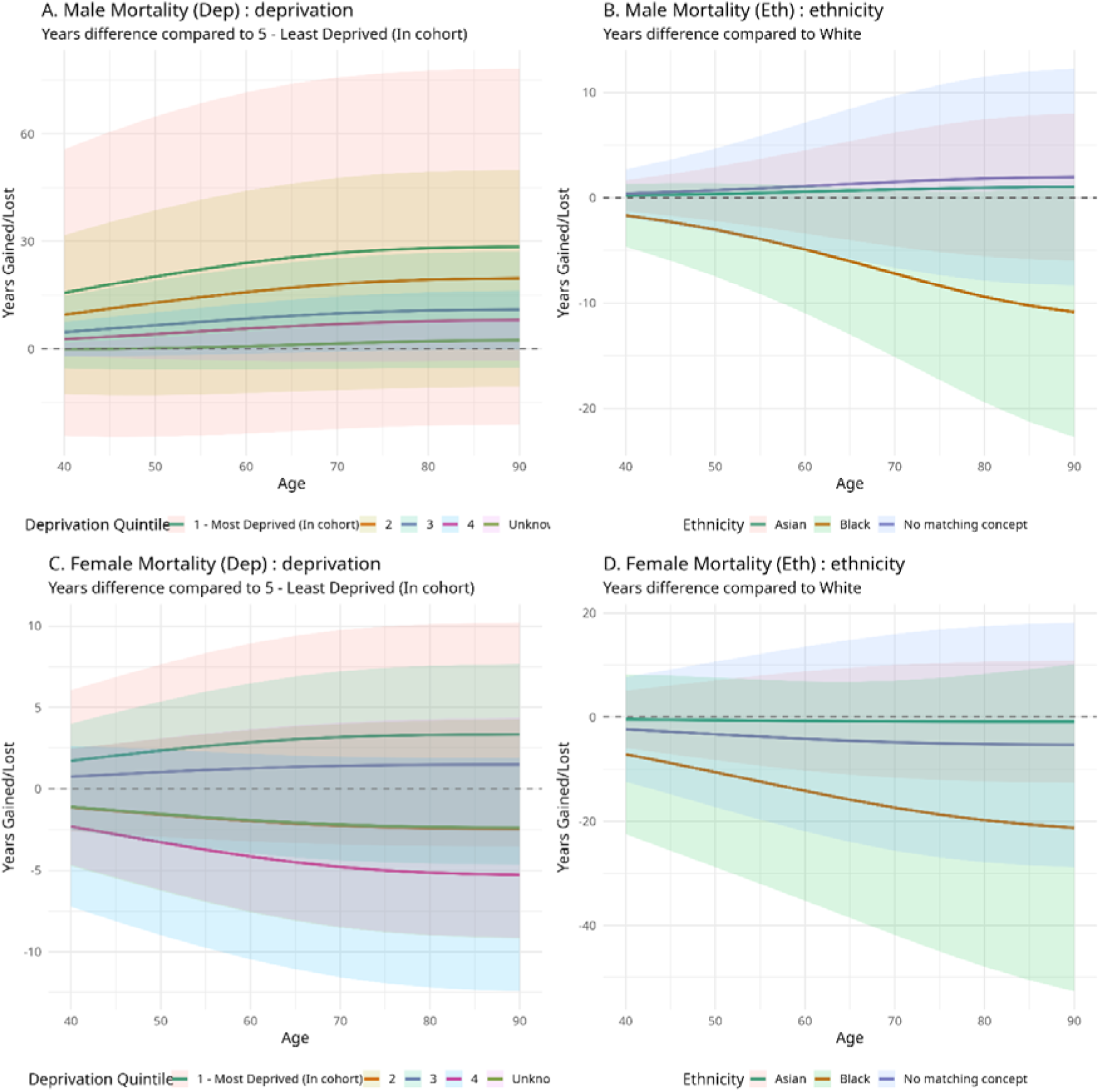
Differences in RMST for age at death: Years gained or lost. Note: Positive values on the Y-axis indicate greater years lost relative to the reference group

Panel B shows RMST differences for the male cohort by ethnicity, with the White cohort used as the reference category. The Black cohort demonstrates the strongest negative trajectory, indicating progressively fewer years of expected survival relative to White men with increasing age, whereas the Asian cohort remains comparatively close to the reference group.

Panels C and D present the equivalent analyses for females. Compared with males, the female trajectories are noticeably flatter and more stable across age. In particular, the deprivation-related differences in Panel C appear relatively constant over time, supporting the finding that deprivation effects among females are less strongly time-varying than those observed in the male cohort.

### Prognosis

When looking at the survival estimates from the first clinic visit, goodness-of-fit analysis using Cox-Snell residuals indicated that age at presentation required non-linear modelling. Age was therefore modelled using natural cubic splines with two degrees of freedom, which improved model stability. These results identified age as the primary driver of mortality (HR 1.05 per year). While early disease identification is critical for all patients, these findings suggest that a DFU in later life can act as a marker for a terminal event.

The absolute deprivation gap remained clinically significant. While relative hazard ratios for the least deprived group were lower than the most deprived areas (HR 0.73, 95% CI 0.63–0.84, p=0.003), the absolute clinical impact is better contextualized by the 5-year survival probability estimates. Here, survival estimate is 59.1% for the least deprived group versus 48.7% for the most deprived (10.4% difference), as shown in Figure 1c. Notably, although males had an earlier age at first visit, sex was not a significant independent predictor of mortality (HR 1.10, 95% CI 0.97–1.25, p=0.115). This indicates that once age at presentation is accounted for, men do not have a worse prognosis than women.

**Figure 1c:**
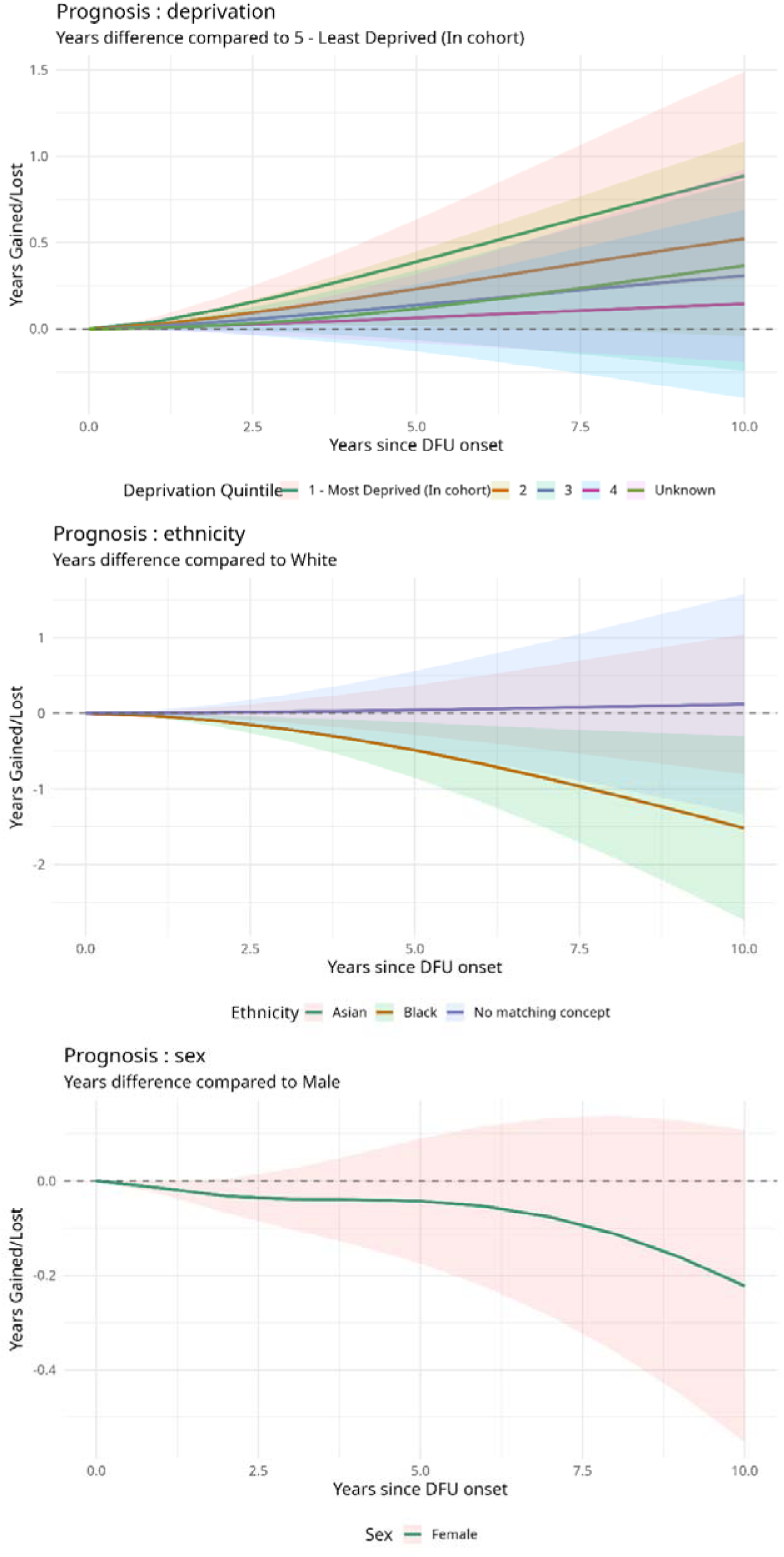
Prognosis RMST Grid (Post-Clinic Survival) Note: Positive values on the Y-axis indicate greater years lost relative to the reference group

Panel A (deprivation) shows a consistent and widening survival gap, with the curves for more deprived quintiles remaining distinctly separated from the reference group (least deprived) across the prognostic horizon. This suggests that the prognostic disadvantage associated with deprivation is sustained over time. Panel B (ethnicity) highlights the relative survival advantage observed in the Black cohort, where the trajectory heads towards years gained compared with the White reference group. Finally, although Panel C (sex) shows a trend toward divergence between male and female prognostic trajectories, this does not reach statistical distinction.

## Discussion

This study uses a registry of active DFU patients linked to the OMOP CDM to provide an investigation of the inequalities inherent in diabetic foot disease onset and mortality. Our key findings are threefold. First, that socioeconomic deprivation acts as a potent accelerator of pathology by shifting the age of DFU onset significantly earlier and reducing post-presentation survival. We show that DFU presentation is 10 years earlier in the most deprived areas, compared to the least deprived areas (table S1). The time-varying effect identified in our analysis, which was most pronounced in middle age, supports the hypothesis that deprivation functions as a cumulative stressor (Geronimus et al., 2006). Our results estimate a 10.4% absolute prognostic survival difference at 5 years from DFU clinic between the most and least deprived groups. This aligns with the hypothesis that deprivation is a fundamental determinant of life expectancy, independent of age and comorbidity burden.

Secondly, our findings refine the traditional view of male disadvantage in diabetic foot disease by identifying where this risk occurs. Male sex was associated with a significant acceleration of disease onset (HR 1.24), meaning men develop DFU earlier in the life course than women. However, this disadvantage did not persist into the clinical prognosis phase. Once age at presentation was accounted for, there was no significant difference in prognostic survival between males and females (HR 1.103, p=0.115).

While a higher prevalence of cardiovascular comorbidity is often cited as a driver of male mortality, adjusting for CCI in our models did not alter these findings. This suggests that prognostically, male disadvantage in DFU is primarily a function of earlier disease onset rather than a worse prognosis following initial clinical presentation.

Thirdly, our results add a complex layer to the ethnicity paradoxes described in the literature (Abbott et al., 2005). Despite small sample size, we observed distinct phenotypic trajectories between minority groups. The Asian cohort presented significantly younger (median age 63 vs 71 for White), consistent with the known earlier onset of Type 2 diabetes in South Asian populations. In contrast, the Black ethnic group presented at an older age (median 73) and with a significantly higher comorbidity burden.

Black patients in this cohort showed a paradoxically lower mortality risk despite being older and having a higher burden of morbidity at their index clinic visit. We believe this is a case of negative confounding. Even though they were typically much sicker at the point of clinical entry, the adjusted model shows these patients lived longer than White counterparts with equivalent morbidity profiles.

This likely reflects a survivor effect driven by structural barriers. The systemic obstacles to specialist care may act as a filter, where only the most physiologically resilient Black patients manage to reach the clinic while navigating advanced disease. Rather than a biological advantage, this result points to a selection bias. The study cohort potentially misses a hidden population of more vulnerable patients who died or remained undiagnosed because they could not overcome the barriers required to access care.

### Strengths and limitations

Our study’s strength lies in using the OMOP CDM to map electronic health records, ensuring reproducible definitions of comorbidities and outcomes. By employing Royston-Parmar flexible parametric models, we moved beyond the limitations of Cox proportional hazards, capturing how drivers of disease fluctuate over time. This approach also handled left truncation, allowing us to visualize life-course survival from birth.

Importantly, our cohort was defined by a clinical DFU photo registration. While this focuses on a care-critical phenotype, it introduces sampling bias because results reflect specialist secondary care and may not represent milder cases managed in primary care. Finally, relative measures like Hazard Ratios were supplemented with RMST to better quantify the absolute mortality gap and life expectancy lost.

These methods enabled important and significant survival trajectories to be identified. These findings warrant further investigation to support the generalisability, especially where group sizes were small.

### Implications

Social determinants must be treated with the same clinical gravity as biological pathology. Shifting from relative risks to absolute survival metrics provides a precise tool for health economic modelling and proactive resource allocation.

The use of the OMOP CDM offers a transformative opportunity to overcome small sample sizes in health equity research. Although current ethnic aggregation may obscure distinct trajectories such as the South Asian paradox, this data standard enables international federation to achieve the statistical power necessary to disaggregate subgroups and identify biosocial-economic phenotypes invisible in single-centre studies.

DFU management is experiencing a digital transition toward AI-driven monitoring and a pharmacological shift toward incretin analogues. While deep learning offers scalable wound classification and drugs like semaglutide reduces metabolic burden, equitable deployment remains precarious. If algorithms use biased datasets or breakthroughs remain inaccessible to deprived populations, these innovations risk widening existing health-equity gaps.

### Conclusion

Diabetic foot ulcers represent a complex convergence of biological pathology and systemic health inequality. The present study demonstrates that socio-economic deprivation significantly accelerates disease onset, reducing DFU-free survival by 13.4% at age 70, and contributes to up to four years of life lost. Following clinical presentation, this disparity manifests as a 10.4% absolute survival deficit at five years for the most deprived cohorts. Furthermore, the analysis identifies distinct demographic risk trajectories. Whilst male sex precipitates significantly earlier disease onset (HR 1.24), post-presentation prognosis is principally driven by age (HR 1.05 per year) rather than sex, positioning later-life incidence as a critical marker of terminal decline. The data also reveal profound structural disparities amongst ethnic minority cohorts, notably a survivor-effect selection bias within Black populations (HR 0.49). By objectively quantifying these disparities, this study highlights the substantial clinical burden of socio-economic determinants. These findings underscore the necessity of transitioning from reactive management towards proactive interventions, utilising robust data governance and technological advancements to facilitate the equitable delivery of precision healthcare.

## Data Availability

All data produced in the present study are covered by the specific project ethics approval, and GDPR of NHS research data. Access is granted via the LSC SDE Data Access Group.

## Supplementary material

## S1: Baseline characteristics by deprivation

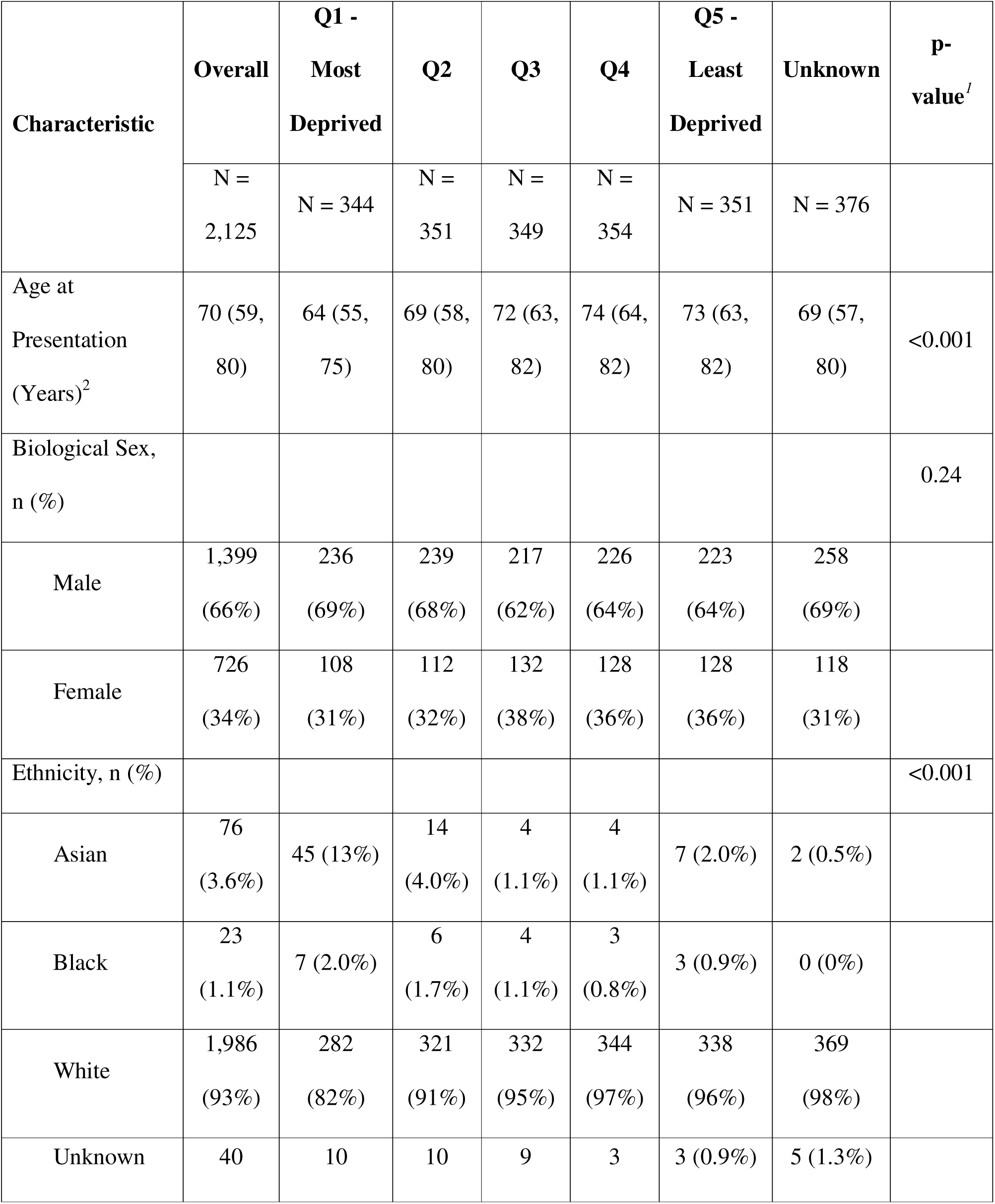
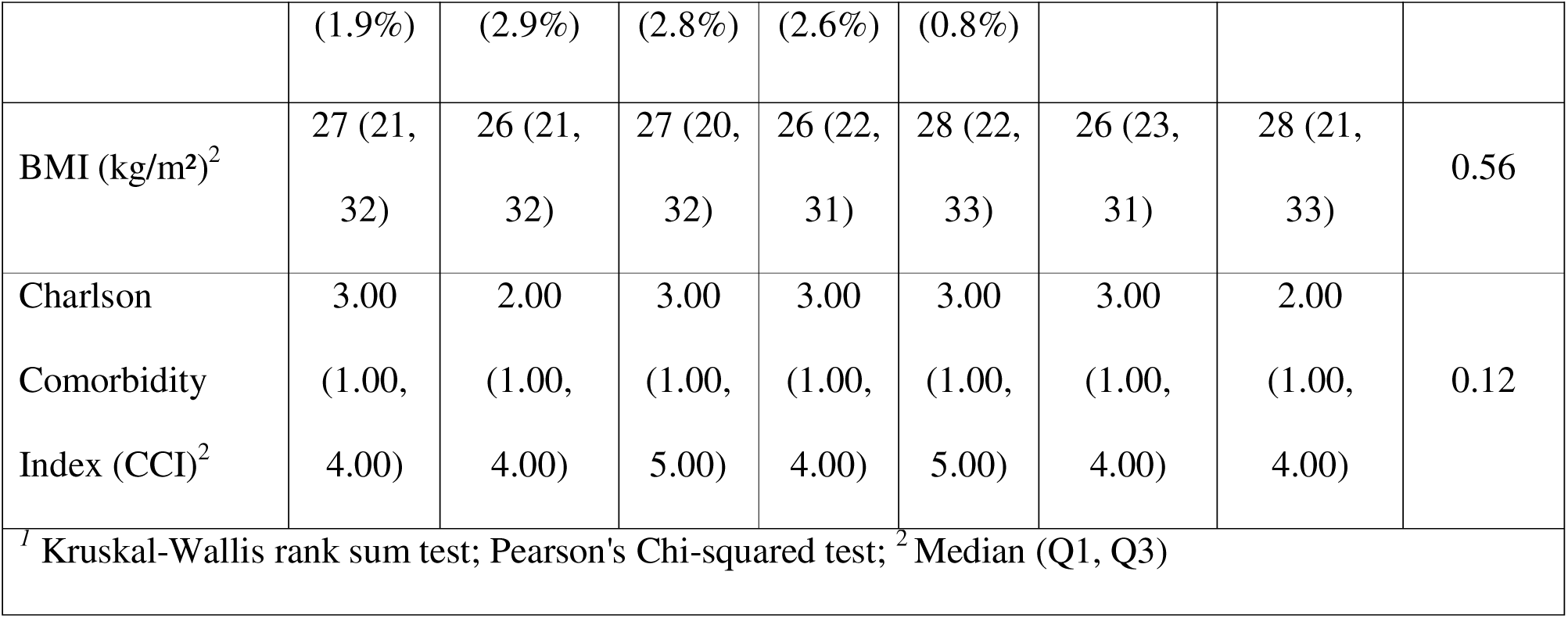

## S2: Baseline characteristics by ethnicity

| Characteristic | Asian | Black | White | Unknown | p-value<sup>1</sup> |
| --- | --- | --- | --- | --- | --- |
| Age at first visit (Years)<sup>2</sup> | 63 (56, 77) | 73 (65, 82) | 71 (60, 80) | 67 (57, 78) | 0.008 |
| Biological Sex, n (%) |  |  |  |  | 0.71 |
| Male | 52 (68) | 17 (74) | 1,302 (66) | 28 (70) |  |
| Female | 24 (32) | 6 (26) | 684 (34) | 12 (30) |  |
| Deprivation Quintile, n (%) |  |  |  |  | <0.001 |
| Q1 - Most Deprived | 45 (59) | 7 (30) | 282 (14) | 10 (25) |  |
| Q2 | 14 (18) | 6 (26) | 321 (16) | 10 (25) |  |
| Q3 | 4 (5.3) | 4 (17) | 332 (17) | 9 (23) |  |
| Q4 | 4 (5.3) | 3 (13) | 344 (17) | 3 (7.5) |  |
| Q5 - Least Deprived | 7 (9.2) | 3 (13) | 338 (17) | 3 (7.5) |  |
| Body Mass Index (kg/m<sup>2</sup>) | 26 (22, 31) | 31 (25, 41) | 27 (22, 32) | 23 (20, 27) | 0.075 |
| Charlson Comorbidity Index (CCI) | 3.00 (1.00, 4.50) | 3.00 (1.00, 5.00) | 3.00 (1.00, 4.00) | 1.00 (0.00, 2.50) | 0.001 |
| <sup>1</sup> Kruskal-Wallis rank sum test; Pearson's Chi-squared test; <sup>2</sup> Median (Q1, Q3) |  |  |  |  |  |

S3: FPM Survival Probability by Deprivation

## S3: FPM Survival Probability by Deprivation

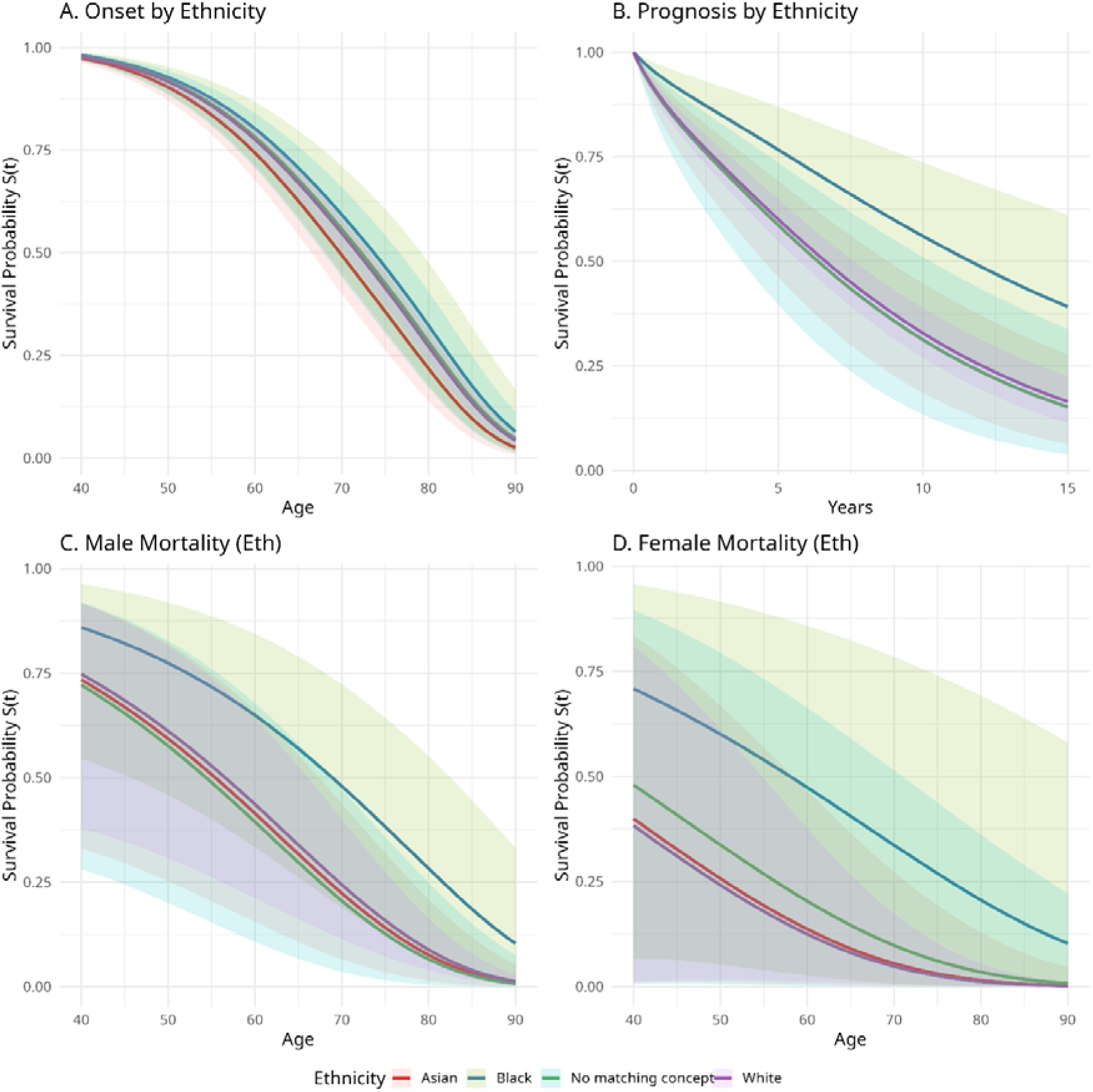

## S4: FPM Survival Probability by Ethnicity

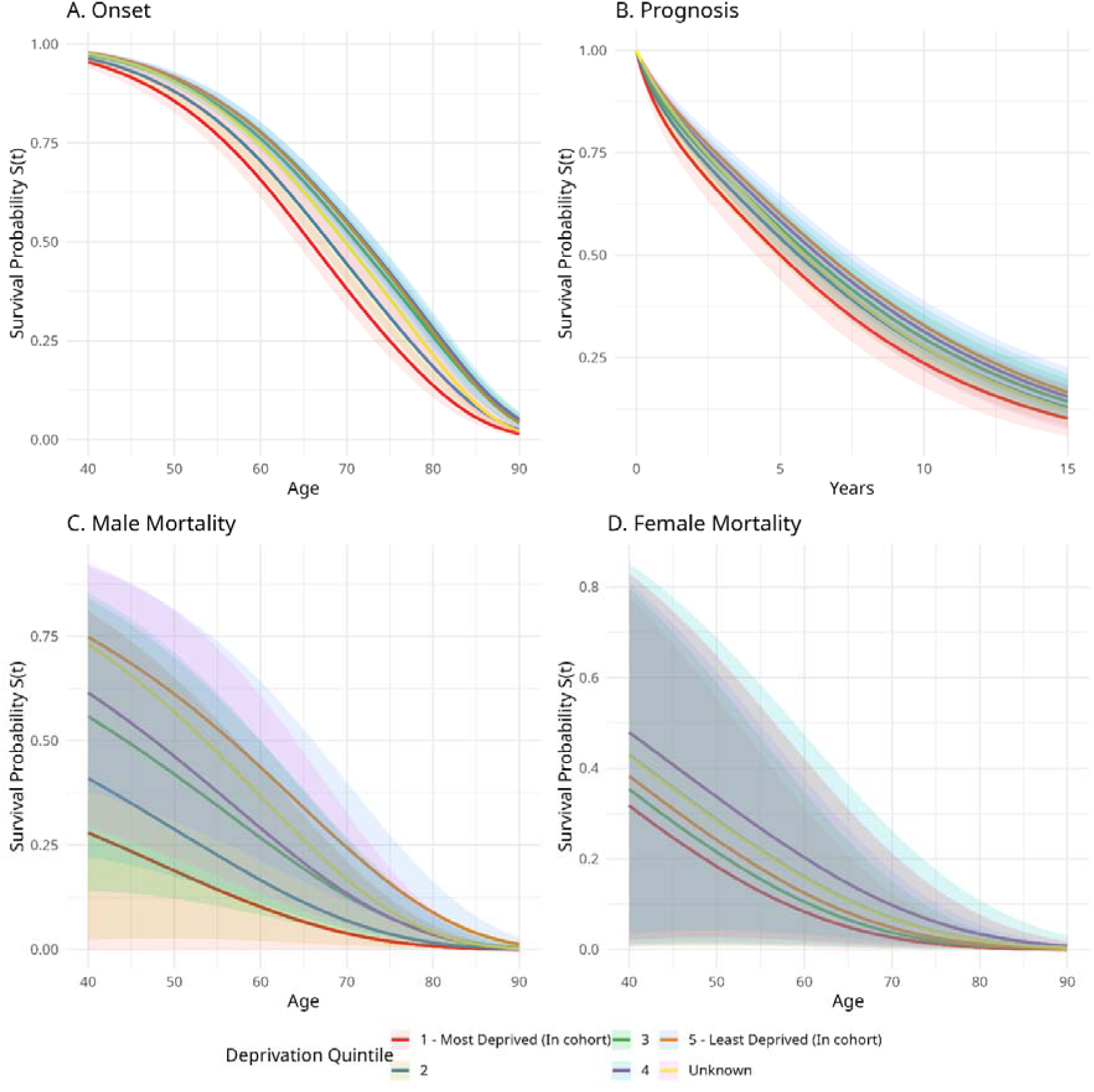

## Notes

### Competing Interest Statement

The authors have declared no competing interest.

### Author Declarations

Ethical approval for this study was obtained from the NHS Research Ethics Committee (REC reference: 15/NW/0539; IRAS project ID: 159863).

